# Examining Brain Structures and Cognitive Functions in Patients with Recovered COVID-19 Infection: A Multicenter Study Using 7T MRI

**DOI:** 10.1101/2024.11.13.24317121

**Authors:** Jr-Jiun Liou, Tales Santini, Jinghang Li, Monica Gireud-Goss, Vibhuti Patel, Oluwatobi F. Adeyemi, Gabriel A. de Erausquin, Valentina R. Garbarino, Mohamad Habes, Jayandra J. Himali, Christof Karmonik, Beth E. Snitz, Joseph M. Mettenburg, Minjie Wu, Howard J. Aizenstein, Anna L. Marsland, Peter J. Gianaros, Richard Bowtell, Olivier Mougin, Farhaan S. Vahidy, Timothy D. Girard, Heidi I.L. Jacobs, Akram A. Hosseini, Sudha Seshadri, Tamer S. Ibrahim

## Abstract

**Importance:** Emerging evidence suggests that severe acute respiratory syndrome, COVID-19, negatively impacts brain health, with clinical magnetic resonance imaging (MRI) showing a wide range of neurologic manifestations but no consistent pattern. Compared with 3 Tesla (3T) MRI, 7 Tesla (7T) MRI can detect more subtle injuries, including hippocampal subfield volume differences and additional standard biomarkers such as white matter lesions. 7T MRI could help with the interpretation of the various persistent post-acute and distal onset sequelae of COVID-19 infection.

**Objective:** To investigate the differences in white matter hyperintensity (WMH), hippocampal subfields volumes, and cognition between patients hospitalized with COVID-19 and non-hospitalized participants in a multi-site/multi-national cohort.

**Design:** Original investigation of patients hospitalized with COVID-19 between 5/2020 and 10/2022 in 3 USA and 1 UK medical centers with follow-up at hospital discharge.

**Participants:** A total of 179 participants without a history of dementia completed cognitive, mood and other assessments and MRI scans.

**Exposure:** COVID-19 severity, as measured by hospitalization vs no hospitalization

**Main Outcomes and Measures:** 7T MRI scans were acquired. All WMH and hippocampal subfield volumes were corrected for intracranial volumes to account for subject variability. Cognition was assessed using a comprehensive battery of tests. Pearson correlations and unpaired t-tests were performed to assess correlations and differences between hospitalized and non-hospitalized groups.

**Results:** We found similar WMH volume (4112 vs 3144mm³, p=0.2131), smaller hippocampal volume (11856 vs 12227mm³, p=0.0497) and lower cognitive and memory performance, especially the MoCA score (24.9 vs 26.4 pts, p=0.0084), duration completing trail making test B (97.6 vs 79.4 seconds, p=0.0285), Craft immediate recall (12.6 vs 16.4 pts, p<0.0001), Craft delay recall (12.0 vs 15.6 pts, p=0.0001), and Benson figure copy (15.2 vs 16.1 pts, p=0.0078) in 52 patients hospitalized for COVID-19 (19[37%] female; mean[SD] age, 61.1[7.4] years) compared with 111 age-matched non-hospitalized participants (66[59%] female; mean[SD] age, 61.5[8.4] years).

**Conclusions and Relevance:** Our results indicate that hospitalized COVID-19 cases show lower hippocampal volume when compared to non-hospitalized participants. We also show that WMH and hippocampal volumes correlate with worse cognitive scores in hospitalized patients compared with non-hospitalized participants, potentially indicating recent lesions and atrophy.

**Key Points:** Question: Do white matter hyperintensity burden, hippocampal whole and subfield volumes, and cognition differ between patients hospitalized with COVID-19 versus participants without hospitalization?

Findings: We found no significant difference in white matter hyperintensity volume, but hippocampal volume was reduced, and cognitive and memory performance were worse in those hospitalized for COVID-19 compared with age-matched non-hospitalized group (either mild COVID-19 or no COVID-19 reported). In the hospitalized group, increased white matter hyperintensity and reduced hippocampal volumes are significantly higher correlated with worse cognitive and memory scores.

Meaning: Adults hospitalized for COVID-19 had lower hippocampal volumes and worse cognitive performance than adults with COVID-19 that did not lead to hospitalization or without reported COVID-19 infection.

## Introduction

Emerging evidence indicates that COVID-19 can negatively impact brain health ^1,2^. However, clinical magnetic resonance imaging (MRI) reveals a wide range of neurologic manifestations without a consistent pattern ^3–7^. A recent observational cohort in the United Kingdom found that COVID-19 patients have more white matter hyperintensities (WMHs) and reduced regional brain volume detected using 3 Tesla (3T) MRI compared to controls ^8^. Whole hippocampal atrophy, linked to cognitive and memory decline, has been noted in patients with post-acute sequelae of COVID-19 infection (PASC) ^9,10^. Aged individuals after a COVID-19 diagnosis are at higher risk for new diagnoses of Alzheimer’s disease (AD) within a year of infection. ^11^ Further, plasma biomarker levels associated with neurodegeneration and AD (i.e. GFAP, NfL) are elevated in non-demented COVID patients when compared to individuals with AD who have not experienced a COVID-19 infection ^12^.

Given that PASC is associated with persistent, relapsing, and new symptoms, characterizing PASC in a diverse prospective observational cohort could be beneficial to improving treatment and prevention of these COVID-19 associated complications ^13–17^. 7 Tesla (7T) MRI can detect more subtle injury in the brain when compared with the more available MRIs at 1.5T and 3T. 7T detects 5% more white matter lesions ^18^, 42% more central vessels ^18^, and three times more cerebral microinfarcts ^19^, suggesting a much higher detection rate at 7T. Using 7T MRI, we can detect differences between specific and nonspecific white matter hyperintensities ^20^, more subtle perivascular spaces ^21^, and nuanced changes in hippocampal subfields ^22,23^.

The purpose of this study is to investigate the association of COVID-19 hospitalization for COVID-19 with WMH burden, hippocampal volume, and cognitive measures within a diverse multinational cohort from the 7T MRI Covid Consortium.

## Methods

### Participant recruitment

With Institutional Review Board approval from the University of Pittsburgh, Houston Methodist Research Institute, the University of Texas Health San Antonio, and the University of Nottingham, participants were identified either from outreach referrals, clinical referrals, or inpatient hospital records at four locations: Pittsburgh, USA; Houston, USA; San Antonio, USA; and Nottingham, UK. Prior to their initial visit, participants underwent a comprehensive informed consent process, which included a detailed review of the study’s objectives. Eligible participants were between the ages of 45 and 85 and had no pre-existing dementia, as screened using the Informant Questionnaire on Cognitive Decline in the Elderly and the Clinical Dementia Rating scale. Participants were asked to participate in two visits, the first to capture medical background, and completion of physical and cognitive assessments, and a second to capture a 7T MRI. A total of 179 participants completed 7T MRI scans, of which 163 were acquired with acceptable image quality for inclusion in our analysis **(Figure 1)**.

**Figure 1.**
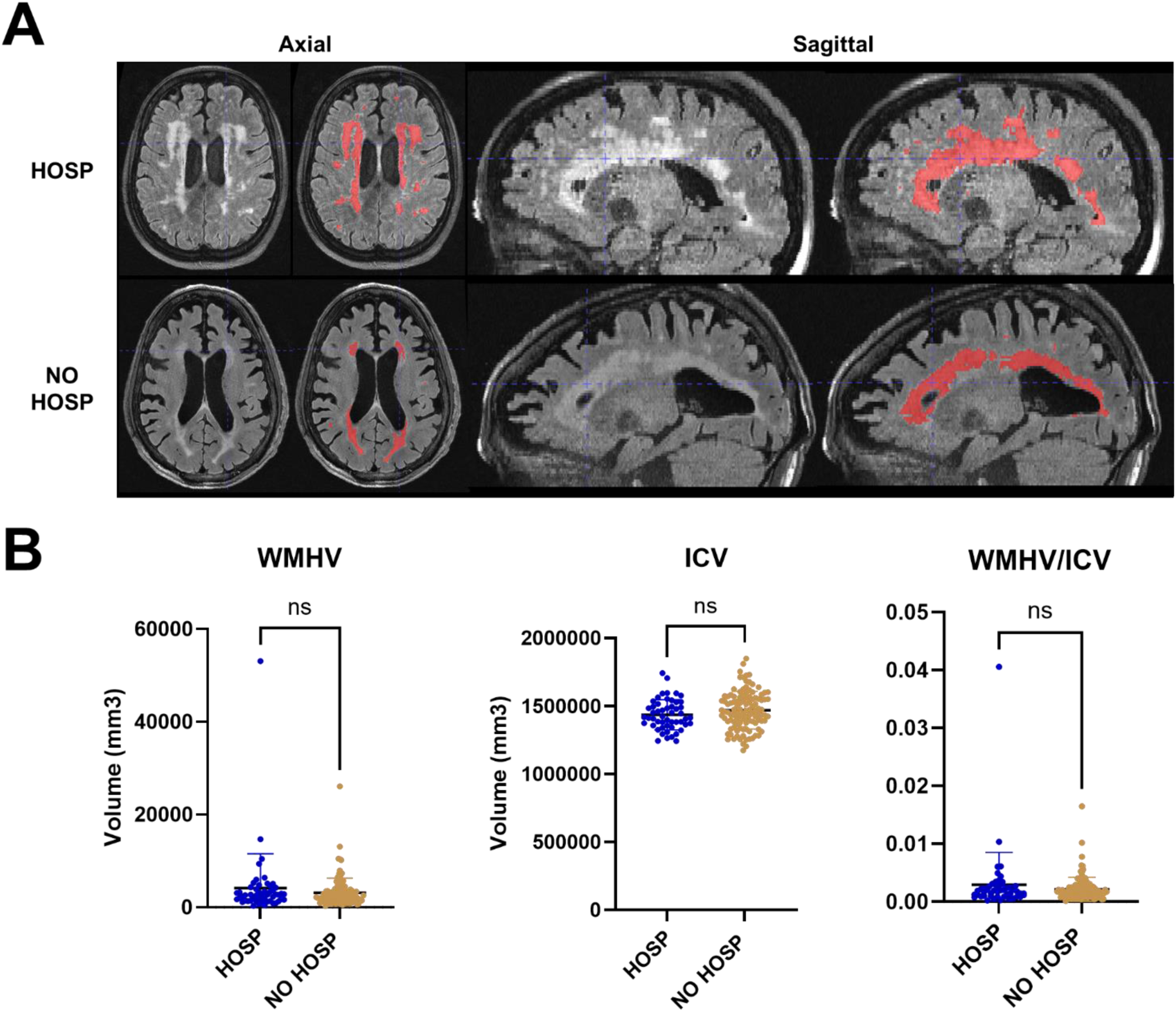
White matter hyperintensity segmentation and quantification. (A) Representative T2-FLAIR images, with and without segmentation, from two participants with the highest WMH burden in each group in both axial and sagittal views, demonstrate a higher WMH burden in the HOSP group. (B) At the group level, no significant difference was detected in total WMH volume. Error bars represent standard deviations.

### COVID-19 severity

The 163 participants with acceptable quality MRIs were categorized into two groups: 52 participants who were hospitalized for COVID-19 (HOSP, which included participants hospitalized for suspected or diagnosed with COVID-19 prior to the 7T MRI visit) and 111 participants who were not hospitalized (NO HOSP, which included mild and not-reported COVID-19 infection in the last 6-15 months prior to 7T MRI the visit). According to the WHO Clinical Classification, the severity of acute COVID-19 illness is classified as Mild, Moderate, Severe, and Critical, and each participant was also classified into one of these categories as applicable based on their reported symptoms. **(Table S1)**.

### Demographic information

Demographic information, lifestyle habits, vascular risk factors, medication usage, and medical history data were collected. Categories for years of education correspond to completion of high school (12), professional/trade school (14), bachelor’s degree (16), master’s degree (18), or doctorate (20). Demographic assessments encompass age, biological sex, race, and ethnicity, as well as occupational history utilizing a PhenX tool, and information on current and former smoking, alcohol, and recreational drug use.

### Medical history

Key responses in medical, cardiovascular, and neurological conditions were included: diabetes, hypertension, hypercholesterolemia, cancer, B12 deficiency, thyroid disease, sleep apnea, obesity (BMI>30), atrial fibrillation, congestive heart failure, heart valve replacement or repair, migraine, and head injury. Detailed records of prior COVID-19 illness and vaccination history were also documented. This included whether the participant received a COVID-19 vaccine, the number of doses, the product name, and the date of each dose. Additionally, flu vaccination history and the date of flu vaccination were recorded.

### Cognitive, mood and other neuropsychological assessments

Comprehensive evaluations encompassing cognitive, neuropsychological, mood, and functional domains were conducted. The assessment battery comprises the Uniform Data Set neuropsychological test battery from the National Alzheimer’s Coordinating Center (NACC) and NIH Toolbox. The cognitive test battery assesses verbal memory (Craft Story immediate/delayed recall), visual memory (Benson Complex Figure intermediate/delayed recall), psychomotor speed and cognitive flexibility (Trail Making Test), and visuospatial construction (Benson Figure copy).

### Neuroimaging

During the second visit, each participant completed a 75-minute MRI scan. Participants in Pittsburgh ^24^ were imaged using a 7T Siemens MAGNETOM human MRI scanner (Erlangen, Germany) equipped with the 2^nd^ ^28^ generation of the radiofrequency Tic-Tac-Toe head coil at University of Pittsburgh. Participants in Houston and San Antonio ^29^ were imaged using a 7T Siemens MAGNETOM Terra (Erlangen, Germany) at the Houston Methodist Hospital Research Institute with the commercial head coil (NOVA medical) whereas participants in Nottingham ^30^ were imaged using a 7T Philips Achieva (Amsterdam, Netherlands), also using the commercial RF coil (NOVA medical) at the University of Nottingham. Whole brain structural imaging encompassed the acquisition of both T1-MP2RAGE images at a resolution of 0.55 mm isotropic and T2-FLAIR images at a resolution of 0.75×0.75×1.5 mm^3^. A T2-TSE sequence at resolution of 0.375×0.375×1.5mm^3^ was obtained for hippocampal subfield segmentation, acquired perpendicular to the main axis of the hippocampus. The comprehensive sequence parameters are provided in **Table S2**. The MRI scans were performed at an average of 19 months (19.0 ± 7.5 months) after hospital discharge.

### Image analysis

Intracranial volumes were estimated from T1-MP2RAGE using SynthStrip ^31^, which leverages deep learning techniques for improved accuracy. Precise estimation of intracranial volume is crucial for normalizing subregional volumes to account for individual head size variability.

Subsequently, segmentation of WMH from the T2-FLAIR images was performed employing our in-house deep learning-based model wmh_seg2d ^32^. Specifically, using a Unet architecture, the T2-FLAIR images went through four sets of overlap patch embedding and transformer block for encoding and convolutional decoder blocks for decoding. This model, which was trained in a diverse dataset and using data augmentation for the common artifacts seen in 7T MRI, was utilized in the T2-FLAIR images and provides a more accurate WMH segmentation compared to FreeSurfer segmentation of T1w images qualitatively ^32^. All segmentations underwent visual inspection and manual correction by two authors (JJL and TS) using ITK-SNAP ^33^ before proceeding to volume extraction and statistical analysis.

For segmentation of the hippocampal subfields, we first removed the characteristic background noise from the T1-MP2RAGE by combining the white matter, grey matter, cerebrospinal fluid, and skull segmentations from the second inversion of the T1-MP2RAGE sequence obtained using Statistical Parametric Mapping (SPM) 12 package ^34^ and masking the T1-MP2RAGE. We performed an image harmonization between our MRI acquisitions and the atlas in two steps. First, the T2-TSE images were denoised using a variance stabilizing transformation and the Block-matching and 4D filtering algorithm ^22,35^ and the resultant image was bias corrected using SPM12. Second, we denoised the T2-TSE images from “IKND Magdeburg Young Adult 7T ^36^” atlas using the same strategy as applied to our T2-TSE images. We then estimated the volumes of hippocampal subfields using the Automatic Segmentation of Hippocampal Subfields (ASHS) package ^37^. The segmentations were then manually verified and corrected following the Berron’17 protocol ^36^. In the main analysis, we combined the dentate gyrus (DG), cornu Ammonis (CA) 2, and CA3 subfields due to the characteristic small volumes of the CA2 and CA3 subfields and, therefore, potentially higher variability of labeling.

### Statistical analysis

The volumes of WMH and hippocampus subfields are corrected by the intracranial volume and age to adjust for subject variability and aging effects, respectively. Two sample t-tests are employed to compare the HOSP and NO HOSP groups. Pearson correlations are used to see the associations of total and subfield volumes in the hippocampus with age and cognitive/memory scores in HOSP and NO HOSP groups.

## Results

### COVID-19 severity

In HOSP group **(Table 1)**, the average duration of hospitalization is 12.3 ± 14.7 days (range 1-76 days) and 18 out of 52 participants were in an intensive care unit for 9.9 ± 7.4 days (range 2-28 days). According to the WHO severity criteria, we encoded unknown, mild, moderate, severe, and critical to 0, 1, 2, 3, and 4. The average disease severity in HOSP is significantly higher than that of NO HOSP (p<0.001). Based on WHO classification, HOSP participants were classified into Critical (27%), Severe (54%), Moderate (13%), Mild (6%). In contrast, NO HOSP participants included 0% Critical, 3% Severe, 6% Moderate, 38% Mild, and the remaining 53% were asymptomatic. Although no difference between HOSP and NO HOSP is detected in the percentage of participants who received a COVID-19 vaccine(s) (p=0.2294), number of doses received (p=0.7597), or history of flu vaccine(s) (p=0.6613), we are unable to report if the participant received a vaccine before or after infection and/or hospitalization. All data were collected 22.4 ± 8.8 months after the infection and/or hospitalization.

**Table 1.** Difference of COVID-19 severity, demographic information, medical history, cognition, and magnetic resonance imaging (MRI) characteristics of 163 participants in hospitalization (HOSP) and without hospitalization (NO HOSP).

|  | HOSP (n=52) | NO HOSP (n=111) | p-value |
| --- | --- | --- | --- |
| COVID-19 |  |  |  |
| • COVID-19 severity |  |  |  |
| ○ Critical, No. (%); mean LOS | 14 (27); 23.8 days | 0 | **<0.0001** |
| ○ Severe, No. (%); mean LOS | 28 (54); 10.7 days | 3 (3) |  |
| ○ Moderate, No. (%); mean LOS | 7 (13); 1.7 days | 7 (6) |  |
| ○ Mild, No. (%); mean LOS | 3 (6); 1.7 days | 42 (38) |  |
| ○ No reported symptoms, No. (%) | 0 | 59 (53) |  |
| • COVID-19 vaccine, No. (%) | 44 (85) | 76 (86) | 0.2294 |
| • COVID-19 vaccine doses, mean (SD) | 3.0 (0.8) | 3.0 (0.6) | 0.7597 |
| • Flu vaccine, No. (%) | 31 (60) | 52 (59) | 0.6613 |
| Demographic information |  |  |  |
| • Age, mean (SD) | 61.1 (7.4) | 61.5 (8.4) | 0.7402 |
| • Sex female, No. (%) | 19 (37) | 66 (59) | **0.0033** |
| • Race white, No. (%) | 36 (69) | 89 (80) | 0.0595 |
| • Ethnicity Hispanic/Latino, No. (%) | 7 (13) | 19 (17) | 0.5259 |
| • Years of education, mean (SD) | 14.8 (2.7) | 15.8 (2.4) | **0.0205** |
| Medical history |  |  |  |
| • Diabetes, No. (%)<sup>a</sup> | 17 (33) | 10 (11) | **0.0021** |
| • Hypertension, No. (%)<sup>a</sup> | 23 (44) | 30 (34) | 0.2773 |
| • Hypercholesterolemia, No. (%)<sup>a</sup> | 22 (42) | 32 (37) | 0.3843 |
| • Cancer, No. (%)<sup>a</sup> | 8 (15) | 15 (17) | 0.7110 |
| • B12 deficiency, No. (%)<sup>a</sup> | 7 (13) | 13 (15) | 0.8252 |
| • Thyroid disease, No. (%)<sup>a</sup> | 4 (8) | 17 (20) | **0.0496** |
| • Sleep apnea, No. (%)<sup>a</sup> | 13 (25) | 9 (10) | **0.0191** |
| • Obesity (BMI>30), No. (%)<sup>a</sup> | 19 (37) | 16 (18) | **0.0174** |
| • Atrial fibrillation, No. (%)<sup>a</sup> | 3 (6) | 3 (3) | 0.5476 |
| • Congestive heart failure, No. (%)<sup>a</sup> | 1 (2) | 1 (1) | 0.7260 |
| • Heart valve surgery, No. (%)<sup>a</sup> | 1 (2) | 0 | 0.2022 |
| • Migraines, No. (%)<sup>a</sup> | 8 (15) | 21 (24) | 0.1977 |
| • Head Injury, No. (%)<sup>a</sup> | 5 (10) | 16 (18) | 0.1411 |
| Cognition |  |  |  |
| • MoCA, mean (SD) | 24.9 (3.7) | 26.4 (2.9) | **0.0084** |
| • Trail making test A, mean (SD) | 35.0 (22.5) | 32.7 (14.3) | 0.4466 |
| • Trail making test B, mean (SD) | 97.6 (46.0) | 79.4 (48.3) | **0.0285** |
| • Craft immediate recall, mean (SD)<sup>a</sup> | 12.6 (6.4) | 16.4 (3.2) | **<0.0001** |
| • Craft delayed recall, mean (SD)<sup>a</sup> | 12.0 (6.6) | 15.6 (3.7) | **0.0001** |
| • Benson figure copy, mean (SD)<sup>a</sup> | 15.2 (2.7) | 16.1 (1.0) | **0.0078** |
| • Benson figure recall, mean (SD)<sup>a</sup> | 12.1 (3.6) | 12.6 (2.4) | 0.3179 |
| MRI Neuroimaging |  |  |  |
| • ICV, mean (SD) | 1435854 (111163) | 1469898 (138349) | 0.1221 |
| • WMHV, mean (SD) | 4112 (7389) | 3144 (3149) | 0.2420 |
| • WMHV/ICV, mean (SD) | 0.0029 (0.0056) | 0.0021 (0.0021) | 0.2131 |
| • Left+Right Hippocampi, mean (SD) | 11856 (1088) | 12227 (1070) | **0.0497** |
| • Left CA1, mean (SD) | 718 (101) | 745 (116) | 0.0686 |
| • Left CA2+CA3+DG, mean (SD) | 635 (94) | 658 (90) | 0.0684 |
| • Left SUB, mean (SD) | 1073 (138) | 1077 (121) | 0.9442 |
| • Left ERC, mean (SD) | 869 (134) | 873 (127) | 0.9714 |
| • Right CA1, mean (SD) | 755 (98) | 794 (113) | **0.0450** |
| • Right CA2+CA3+DG, mean (SD) | 701 (101) | 722 (87) | 0.1136 |
| • Right SUB, mean (SD) | 1090 (148) | 1111 (124) | 0.5824 |
| • Right ERC, mean (SD) | 876 (122) | 876 (138) | 0.8640 |

Abbreviations: BMI, body mass index; CA1, cornu Ammonis 1; CA2+CA3+DG, cornu Ammonis 2 + cornu Ammonis 3 + dentate gyrus; ERC, entorhinal cortex; ICV, intracranial volume; LOS, length of stay; MoCA, Montreal cognitive assessment; SD, standard deviation; SUB, subiculum; WMHV, white matter hyperintensity volume. <sup>a</sup>Data from 88 participants in NO HOSP group.

### Demographic, medical history, and cognitive/memory performance

The mean age **(Table 1)** of HOSP and NO HOSP is similar - 61.1 ± 7.4 and 61.5 ± 8.4 years, respectively (p=0.7402). No difference was detected in the percentage of self-reported White participants (69% vs 80%, p=0.0595) or Hispanic/Latinos (13% vs 17%, p=0.5259) recruited. Interestingly, the HOSP group was made up of a significantly higher percentage of males (63% vs 41%, p=0.0033) and has fewer years of education than the NO HOSP group (14.8 vs 15.8 years, p=0.0205). The percentages of participants with diabetes (33% vs 11%, p=0.0021), thyroid disease (8% vs 20%, p=0.0496), sleep apnea (25% vs 10%, p=0.0191) and obesity (37% vs 18%, p=0.0174) are significantly higher in HOSP compared to NO HOSP. No significant difference is detected in hypertension, hypercholesterolemia, cancer, B12 deficiency, atrial fibrillation, congestive heart failure, heart valve replacement or repair, migraine, or head injury. In terms of cognitive/memory performance, compared to the NO HOSP group, HOSP participants had significantly worse scores characterized as lower MoCA (24.9 vs 26.4, p=0.0084), higher Trail making test B (97.6 vs 79.4, p=0.0285), lower Craft immediate recall (12.6 vs 16.4, p=<0.0001), lower Craft delayed recall (12.0 vs 16.1, p=0.0001), and lower Benson figure copy (15.2 vs 16.1, p=0.0078) scores. However, no significant difference was detected in Trail Making A (p=0.4466), or Benson figure recall (p=0.3179).

### White matter hyperintensity volume and correlations with cognition

Representative T2-FLAIR images of the two participants with highest WHM burden are presented in axial and sagittal views (Figure 1). HOSP participants have 30% more WMH volume (4112 ± 7389 mm≥ vs 3144 ± 3149 mm≥) compared to the NO HOSP group when controlled for age, however that is not statistically significant (p=0.24) since there is high variability among participants. In the HOSP group, WMH is significantly more associated with worse scores of MoCA (p=0.0006), Trail making test B (p=0.0054), and Benson figure recall (p=0.0117) scores when compared to controls (**Figure 2**).

**Figure 2.**
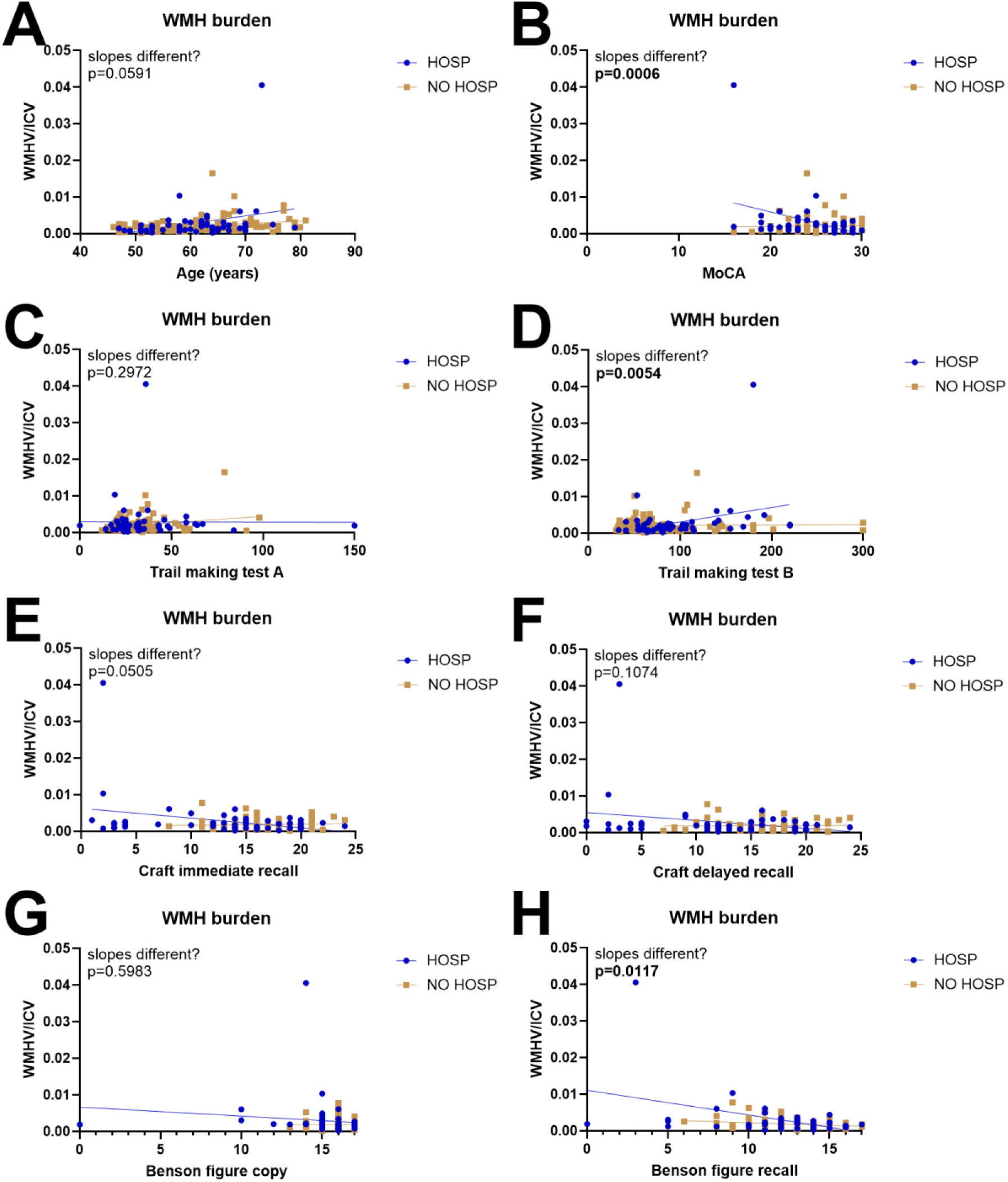
White matter hyperintensity burden and its association with age and cognition. (A) The association with age is similar between groups (p=0.0591). However, the slopes are significantly different between HOSP and NO HOSP in (B) MoCA (p=0.0006), (D) duration to complete Trail making test B (p=0.0054), and (H) the Benson figure recall scores (p=0.0117). The association with (C) duration to complete Trail making test A (p=0.2972), (E) Craft immediate recall (p=0.0505), (F) Craft delayed recall (p=0.1074), and (G) Benson figure copy (p=0.5983) showed no significant differences in slopes between groups. Higher scores are better for MoCA, Craft Immediate Recall, Craft Delayed Recall, Benson Figure Copy, and Benson Figure Recall, and lower scores (shorter times) are better for Trail making Test A and B.

### Hippocampal total and subfield volume

Representative raw, harmonized, and segmented images of hippocampus subfields among the three collection sites – University of Nottingham UK, University of Texas Health San Antonio USA, and University of Pittsburgh USA are presented **(Figure 3).** The dentate gyrus (DG), cornu Ammonis 1, 2, and 3 (CA1, CA2, and CA3), subiculum (Sub), entorhinal cortex (ErC) are labeled in different colors. For quantitative comparisons, the left and right hemispheres are evaluated separately; the DG, CA2, and CA3 are combined given the small volumes of each subfield and risk of mislabeling. The whole hippocampus is significantly smaller in the HOSP group (11856 vs 12227 mm^3^, p=0.0497). Also, the HOSP volumes of Right CA1 (755 vs 794 mm^3^, p=0.045) is significantly lower than the NO HOSP volumes. Although not reaching statistical significance, all other hippocampus subfields show a trend toward smaller volumes in the HOSP group (**Figure 3**). Moreover, the total hippocampal volume is inversely correlated with age, however there is no significant difference in the slope between HOSP and NO HOSP groups (**Figure S3**). In the HOSP groups, larger whole hippocampus is significantly associated with better cognitive/memory scores MoCA (p=0.0119), Craft immediate recall (p=0.0085), and Craft delayed recall (p=0.0053) (**Figure 4**) when compared with the NO HOSP participants. The bilateral subiculum and partially the right DG+CA2+CA3 are driving these findings.

**Figure 3.**
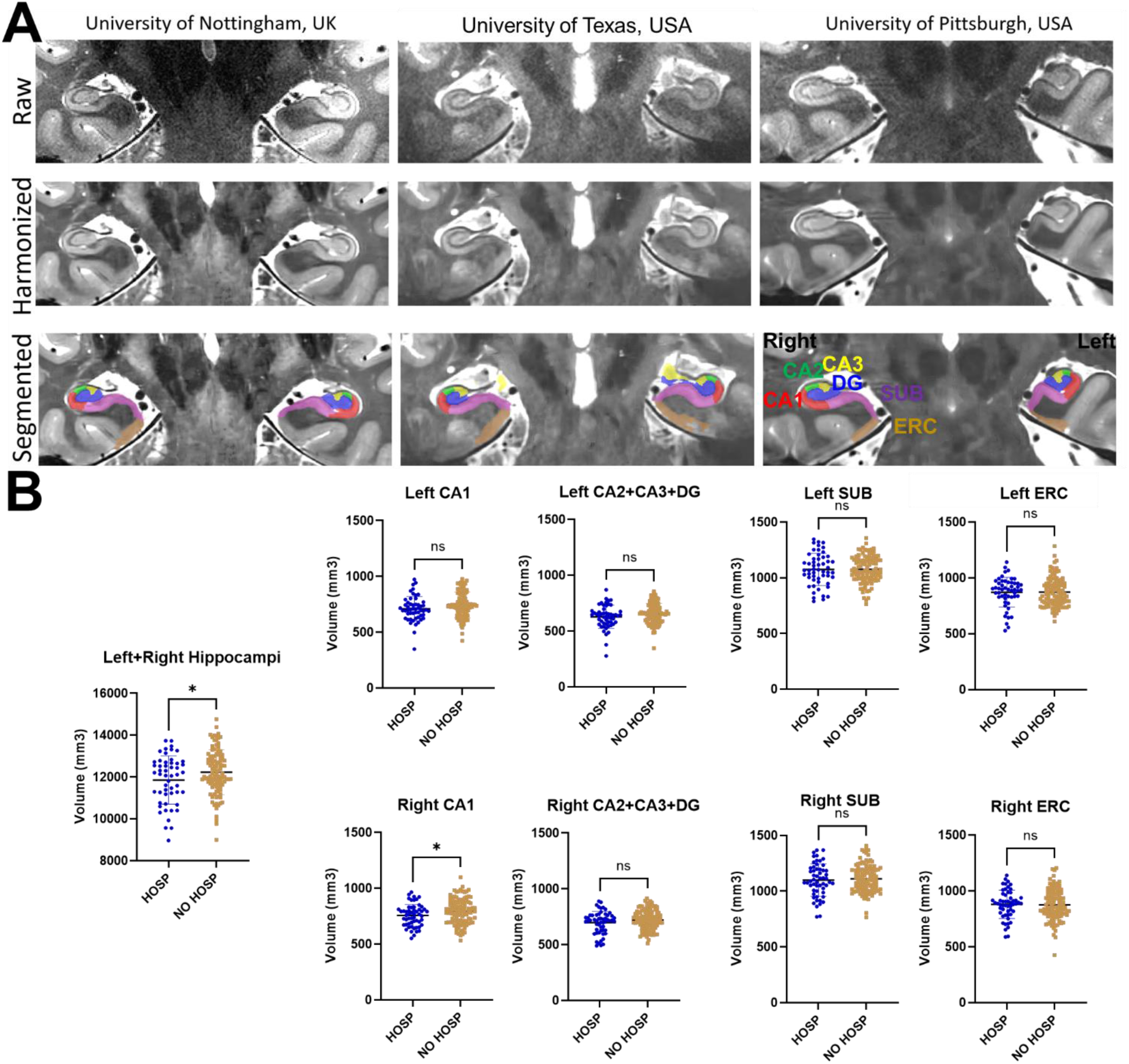
Hippocampal subfield segmentation and quantification. (A) Image harmonization and hippocampal subfield segmentation were performed across three imaging sites. (B) Quantitative comparison of hippocampal subfields between the HOSP and NO HOSP groups revealed significant findings. A reduction in bilateral hippocampal volumes was observed in the HOSP group compared to that of the NO HOSP group (11,856 vs 12,227 mm³, p=0.0497). Additionally, the right CA1 subfield volume was significantly lower in the HOSP group compared to the NO HOSP group (755 vs 794 mm^3^, p=0.045).

**Figure 4.**
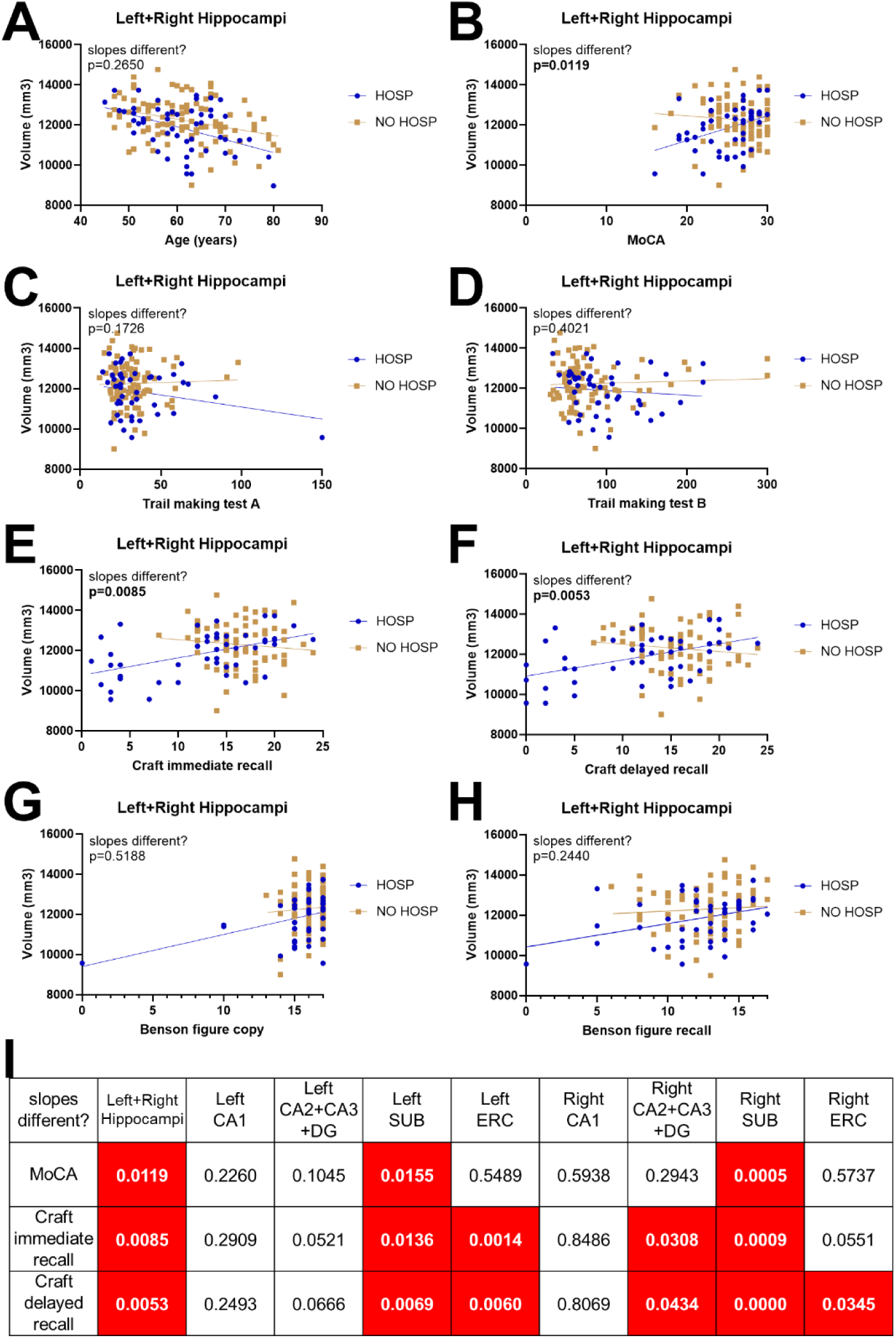
Hippocampal subfield volume and its association with age and cognition. The association of total hippocampal volume with (A) age is similar between groups (p=0.2650). The associations with (B) MoCA (p=0.0119), (E) Craft immediate recall (p=0.0085), and (F) Craft delayed recall (p=0.0053) are significantly different between HOSP and NO HOSP groups. (C-D, G-H) No significant difference between slopes is detected in other cognitive scores: Trail making test A (p=0.1726), Trail making test B (p=0.4021), Benson figure copy (p=0.5188), and Benson figure recall (p=0.2440). (I) Differences in bilateral SUB, bilateral ERC, and right CA2+CA3+DG are also detected. Higher scores are better for MoCA, Craft Immediate Recall, Craft Delayed Recall, Benson Figure Copy, and Benson Figure Recall, and lower scores (shorter times) are better for Trail making Test A and B.

## Discussion

In this study, we investigated the association of hospitalization for COVID-19 with WMH burden and hippocampal volume and cognitive/memory assessments within a diverse multinational cohort using 7T MRI. In the hospitalized group, we found greater associations between higher WMH volumes, lower hippocampus volumes and worse cognitive/memory scores. We also found smaller hippocampal volumes in terms of total and subfields.

WMHs are seen as diffused areas of high intensity on T2-FLAIR images that indicate altered water content in hydrophobic white matter regions ^38^. Several pathological mechanisms underlying WMHs contributions to cognitive impairment and dementia have been proposed: hypoperfusion, defective cerebrovascular reactivity, and blood brain barrier dysfunction as well as dysfunction of oligodendrocyte precursor cells or impaired perivascular clearance ^38^. WMH associations with COVID-19 are mixed in the literature. In larger cohorts of hospitalized patients with COVID-19, 35% ^39^ to 55% ^40^ show nonspecific but likely chronic WMH. When examined in detail, the reported chronic WMH are predominantly located within subcortical and basal ganglia ^39^. However, other studies reported no difference in WMH load between non-hospitalized and hospitalized COVID-19 cases ^41^ or between intensive care unit (ICU) and non-ICU COVID-19 patients ^42^.

Our study showed that COVID-19 patients have a 30% higher WMH burden although the high subject-to-subject variability fails to show statistical significance. Notably, we observed stronger associations between WMHs and three cognitive/memory scores (Figure 2) in the HOSP cohort, potentially suggesting more recent WMHs in the HOSP group and older/chronic WMHs in the NO HOSP group, allowing more time for neuroplastic adaptation. Two potential pathological mechanisms could explain the potentially recent WMH in COVID-19 patients. After COVID-19 infection, COVID-related ischemic stroke, intracranial hemorrhage, and hypoxia brain injury directly result in WMH formation. Alternatively, COVID-19 induces hypoxia and subsequent amyloid beta accumulation in the grey matter, leading to neuronal dysfunction or damage to oligodendrocytes, demyelination and axonal loss in the white matter and ultimately WMH formation. ^43,44^. Also, comorbidities that are higher in the HOSP group (diabetes, sleep apnea, and obesity) may play a role in WMH correlations with cognitive/memory scores. Longitudinal studies are needed to clarify these findings.

Beyond WMH, diffusion-weighted MRI sequences have been shown to be sensitive to white matter microstructural changes; COVID-19 cases show reduced mean diffusivity, axial diffusivity, radial diffusivity and increase of fractional anisotropy in white matter ^10^ along with tissue damage in regions that are functionally connected to the primary olfactory cortex ^1^. Using 7T MRI, other studies have shown that COVID-19 patients exhibit a higher perivascular space count ^45^, enlarged volumes in the pons, superior cerebellar peduncle, and the entire brainstem ^46^ and overall weaker connections based on ROI-to-voxel analysis ^47^ Additionally, significant differences in the brainstem have been observed in post-hospitalization COVID-19 patients ^48^.

Our observation of smaller hippocampal volume is consistent with other studies that found hippocampal volume loss in the COVID-19 group. The severe COVID-19 group showed reduced thickness in the left hippocampus ^49^ when compared to healthy controls. Hippocampal atrophy is accompanied by altered microstructural integrity in white matter and functional connectivity in COVID-19 patients ^9^. COVID-19 survivors have increased amplitude of low-frequency fluctuation values in the hippocampus and parahippocampal gyrus ^50^. A disruption in functional connectivity between the anterior and posterior hippocampus in COVID-19 participants was also observed ^51^. COVID-19 cases showed a higher average regional homogeneity in the right hippocampus compared to healthy controls ^52^. Some studies even extended the investigation into the association between plasma biomarker expression and hippocampal volume. Hospitalized patients have elevated glial fibrillary acidic protein and neurofilament light in the plasma compared to non-hospitalized patients ^9^; the former is associated with volume of the presubiculum and subiculum body in the hippocampus whereas the latter is inversely associated with volume of the hippocampal head subfield ^9^. In contrast, a few studies ^10,53,54^ have shown that COVID-19 patients have enlarged olfactory cortices, bilateral hippocampi, and amygdalae. Additionally, global volume in the bilateral hippocampus correlates with memory loss, and left hippocampal volume is related to smell loss ^10^. This may suggest that acute inflammation leads to edema and observation of hippocampal volume increase, which can be later followed by cell death, tissue degeneration and atrophy.

In our study, we found a lower volume of whole hippocampus and Right CA1 subfield along with a reduced score of MoCA, Craft immediate verbatim, Craft immediate paraphrase, Benson figure copy, and Craft delayed paraphrase in the HOSP group. Moreover, significantly stronger positive associations between hippocampal volumes and better cognitive/memory scores (Figure 4) in the HOSP group may indicate recent atrophy of the hippocampus in this group vs longstanding volumes in the NO HOSP participants. Another explanation would be some comorbidities could be increasing hospitalization rate while also inducing hippocampal atrophy. For instance, hippocampus is especially sensitive to oxidative stress, and sleep apnea could be a factor^55,56^. Longitudinal studies are necessary to elucidate these findings. Moreover, our study shows a slightly and non-significant higher correlation of hippocampus subfields with age in the HOSP group (Figure S3), trending towards significance in the Left CA2+CA3+DG group (p=0.0692), perhaps indicating that older participants in the HOSP group have a larger hippocampal volume gap when compared with the NO HOSP group and that gap decreases in young participants, suggesting that age could be exacerbate the impact of severe COVID-19 on hippocampal volume. Higher sample size is needed to confirm or reject this hypothesis. A hippocampal study identified potential neurobiological mechanisms of central nervous system damage in COVID-19 infection, involving blood brain barrier disruption, elevated levels of interleukins and blunted hippocampal neurogenesis ^57^. Loss of hippocampal neurogenesis may contribute to cognitive and emotional symptoms observed in COVID-19 patients, involving deficits of verbal memory and learning, working memory and executive functions ^57^.

Through autopsy COVID-19 cases, distinct neuropathological features have been identified in human postmortem brain samples. There is significantly higher protein deposition of glial fibrillary acidic protein and C-C motif chemokine 11 and lower protein deposition of myelin oligodendrocyte glycoprotein in the hippocampus of human postmortem tissues ^9^. Autopsy samples from humans with COVID-19 exhibit increased microglial/macrophage reactivity in subcortical white matter compared with cortical grey matter and compared to those without infection ^58^. Significantly higher ionized calcium-binding adapter molecule 1 and interleukin 6 (IL6) signal in the medulla in COVID-19 human brain tissues when compared to control brains, though did not reach significance in the hippocampus ^57^. COVID-19 brains exhibit microglial activation ^59^ and expression of interleukin 1 beta and IL6, especially within the hippocampus and the medulla oblongata, when compared with non-COVID control ^57^. In the hippocampal dentate gyrus of humans infected with COVID-19, fewer neuroblasts and immature neurons are observed ^43,44^. 7T MRI postmortem imaging along with subsequent neuropathological analysis in fatal COVID-19 autopsy cases show a wide range of clinical manifestations but consistent histological expression of reactive gliosis, cortical neuron eosinophilic degeneration, and axonal disruption ^60^. Antemortem-postmortem-histology alignment shows hemorrhage and acute necrosis in the cerebral hemorrhage, suggesting microglial activation, microglial nodules, and neuronophagia observed in most brains are likely due to systemic inflammation, with synergic contribution from hypoxia and ischemia ^61^. COVID-19 causes neuronal degeneration and reduces neurogenesis in human hippocampus ^62^ and that microglial activation is significantly higher in COVID-19 without dementia compared to those without COVID-19 or dementia ^63^.

Our study has several limitations. First, this study is cross-sectional in nature. All HOSP participants completed MRI scans on average 19 months after their COVID-19 hospitalization. Without pre-infection scans, we could not determine whether the observed measures occurred after COVID-19 hospitalization. Longitudinal studies show that COVID-19 patients experience a more dramatic decrease in parahippocampal gyrus tissue integrity and orbitofrontal cortex cortical thickness over a 5 month follow up ^1^. With longitudinal datasets, COVID cases have a greater reduction in grey matter thickness as well as global brain volume and a greater cognitive decline between baseline and five-month follow up ^1^. The adverse effect of COVID-19 on brain function and structure appears to fade over time, especially between ICU and non-ICU patients ^64^. Second, the timing and effect of treatments are not considered in the analysis, although the benefits of hydrocortisone ^65^, benzodiazepine ^66^, anticoagulant heparin ^67^, antiplatelet agents ^68^, and IL6 receptor antagonists ^69^ for the treatment of critically ill patients with COVID-19 have been reported. Timing of the COVID-19 vaccines is also not considered. Participants could have received vaccines prior to COVID-19 infection impacting the presentation of symptom and risk for hospitalization. The third limitation is the analysis of global WMH volumes rather than spatial distributions and regional WMH volumes. Some studies found a higher WMH volume in COVID-19 patients in the right hemisphere ^70^, especially the right frontal lobe ^2^. It is worth noting we only enrolled 4 participants who were hospitalized while testing negative for COVID-19 in the allotted time frame across the study sites. Therefore, the sample size was insufficient to make meaningful comparisons between hospitalization with and without COVID-19 infection.

## Conclusions

In a diverse multinational cohort study using 7T MRI, we found decreased hippocampal volumes as well as reduced cognitive and memory performance in those who were hospitalized for COVID-19 when compared to the age-matched non-hospitalized group. Hospitalized individuals also presented higher associations between higher WMH volume, lower hippocampal volumes and worse cognitive/memory scores.

## Author contributions

Dr. Ibrahim has full access to all the data in the study and takes responsibility for the integrity of the data and the accuracy of the data analysis. Dr. Liou and Dr. Santini contributed equally as co-first authors.

Concept and design: Hosseini, Seshadri, Ibrahim.

Acquisition, analysis, or interpretation of data: Liou, Santini, Li, Goss, Patel, Adeyemi, de Erausquin, Garbarino, Habes, Himali, Karmonik, Li, Masdeu, Snitz, Mettenburg, Wu, Aizenstein, Marsland, Bowtell, Gowland, Roman, Ganguli, Vahidy, Girard, Jacobs, Hosseini, Seshadri, Ibrahim.

Drafting of the manuscript: Liou, Santini, Ibrahim.

Critical revision of the manuscript for important intellectual content: All authors.

Statistical analysis: Liou, Santini.

Obtained funding: Seshadri, Ibrahim.

Administrative, technical, or material support: Goss, Patel, Adeyemi, de Erausquin, Garbarino, Habes, Himali, Karmonik, Li, Masdeu, Snitz, Mettenburg, Wu, Aizenstein, Marsland, Bowtell, Gowland, Roman, Ganguli, Vahidy, Girard, Jacobs, Hosseini, Seshadri, Ibrahim.

Supervision: Hosseini, Seshadri, Ibrahim.

## Acknowledgement

The authors are grateful to all the participants, their families and care providers, and the research and support staff for their contributions to this study. Special thanks to Ms. Caroline J Pidro, Ms. Valerie N Davis, Ms. Janet L Lower, and Mr. Jeremy J. Berardo. We also thank Dr. Vinicius P. Campos and Dr. Marcelo A.C. Vieira for sharing their denoising code.

## Data availability

Demographic and cognitive/memory data as well as the MRI scans will be made available upon reasonable request.

## Supplementary Information

**Figure S1.**
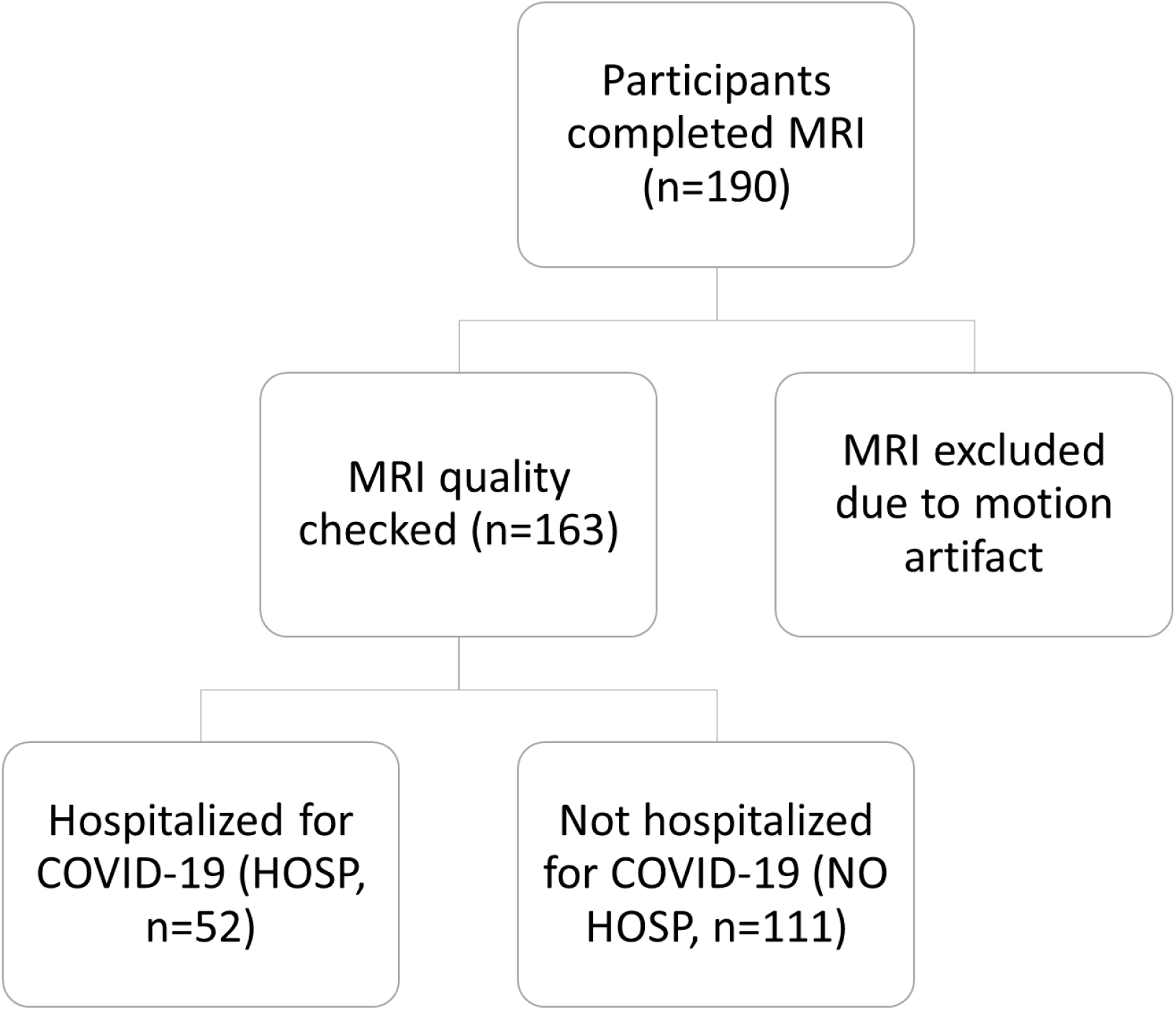
Participant recruitment. Magnetic resonance imaging (MRI) scans from 163 participants were included in our analysis. Participants who were hospitalized for COVID-19 in the past 6-24 months were categorized into the hospitalization (HOSP) group, while the remaining participants were categorized as the non-hospitalization (NO HOSP) group.

**Figure S2.**
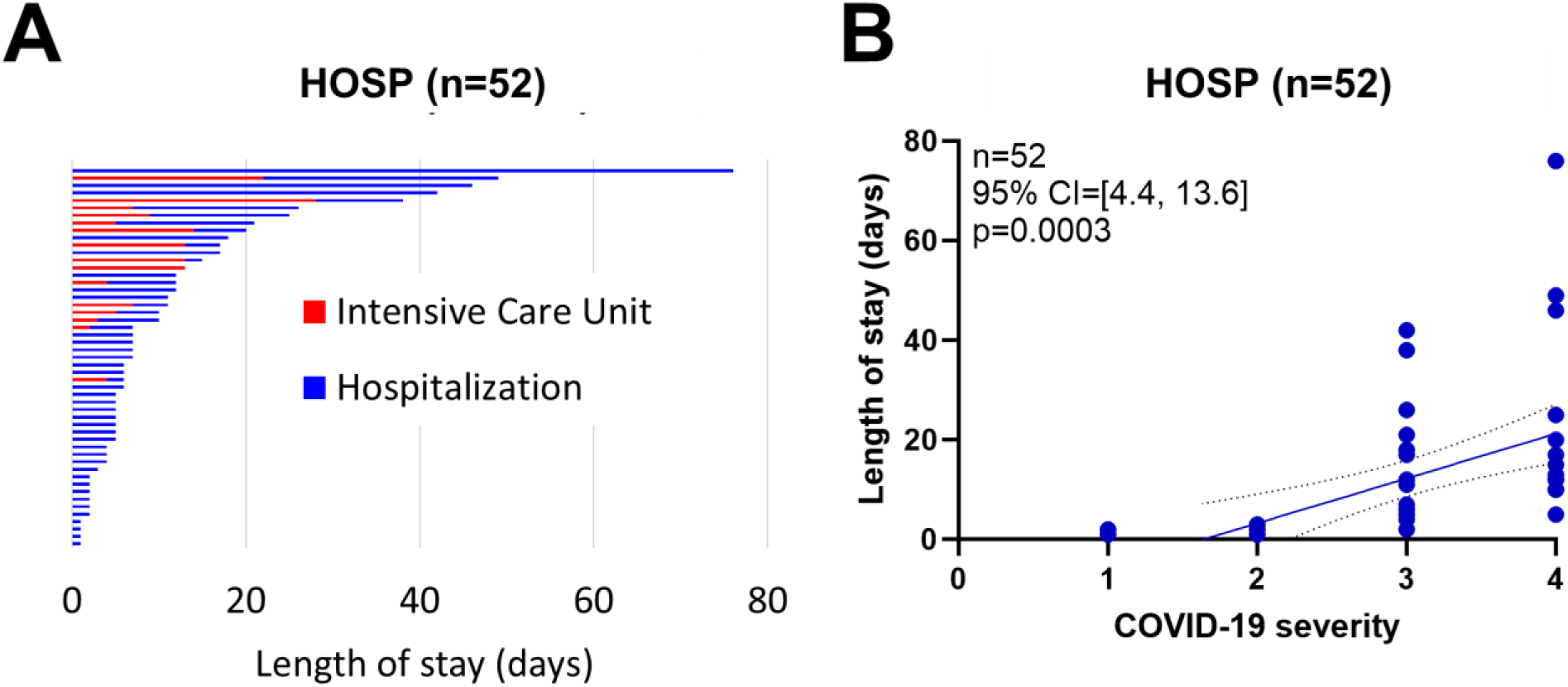
Length of stay and its association with COVID-19 severity in HOSP group. (A) The average duration of hospitalization is 12.3 ± 14.7 days (range 1-76 days) and 18 of those were in an intensive care unit for 9.9 ± 7.4 days (range 2-28 days). (B) According to the WHO classification, we encoded the COVID-19 severity of mild, moderate, severe, and critical to 1, 2, 3, and 4. The length of stay is significantly correlated with COVID-19 severity (p=0.0003).

**Figure S3.**
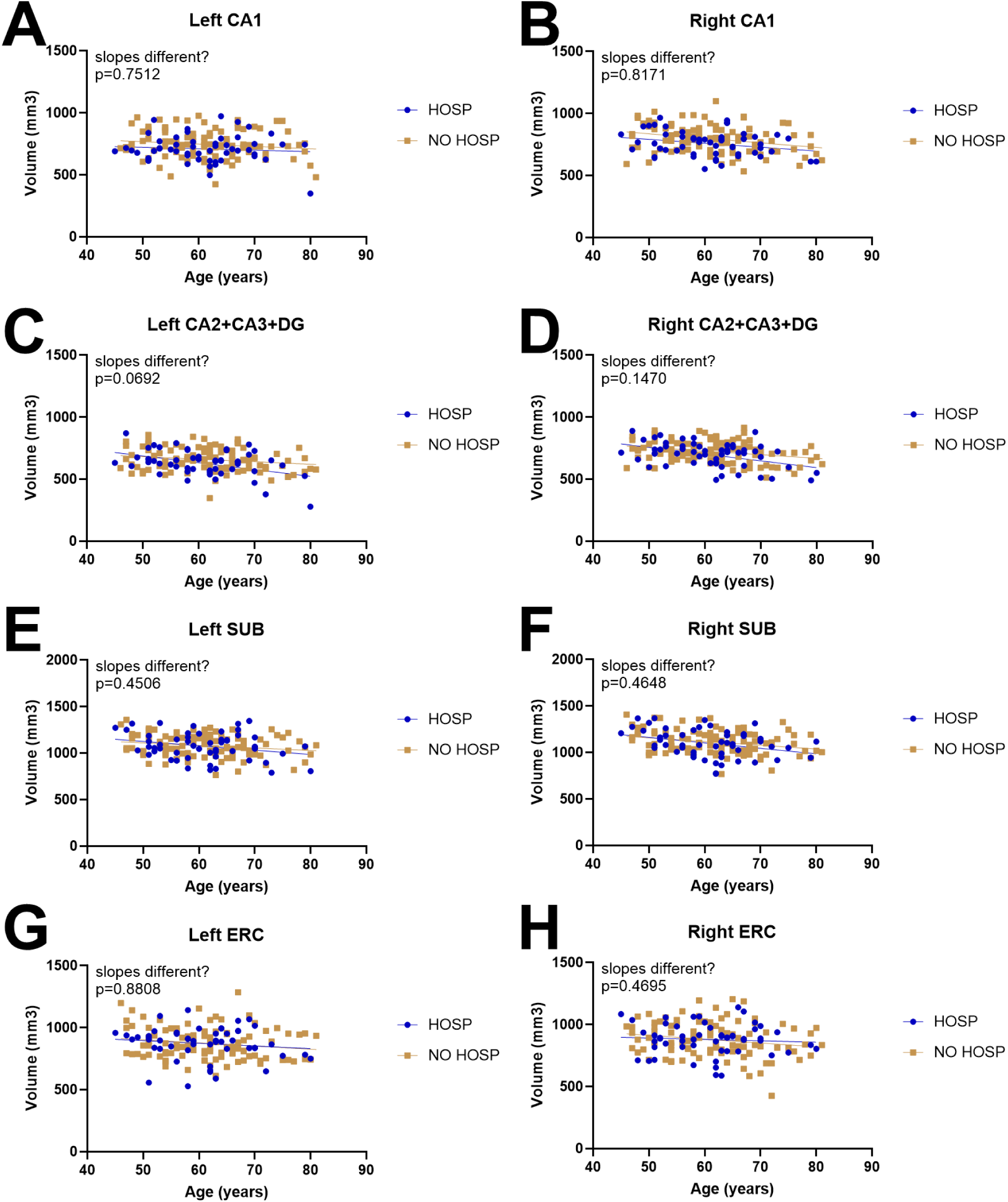
Comparisons of associations of hippocampal subfield volume with age between HOSP and NO HOSP groups. No significant differences in age-related slopes were found between groups for any hippocampal subfields: (A) Left CA1 (p=0.7512), (B) Right CA1 (p=0.8171), (C) Left CA2+CA3+DG (p=0.0692), (D) Right CA2+CA3+DG (p=0.1470), (E) Left SUB (p=0.4506), (F) Right SUB (p=0.4648), (G) Left ERC (p=0.8808), and (H) Right ERC (p=0.4695).

**Table S1.** COVID-19 severity based on WHO classification. SpO2, oxygen saturation rate indicating the oxygen amount being carried by red blood cells in the body which should be equal to or above 96%; RR, respiratory rate which should be about 20 breaths per minute normally; ARDS, acute respiratory distress syndrome.

|  | Based on available clinical records | Based on self-report, if clinical records are not available |
| --- | --- | --- |
| Mild | No hypoxia or pneumonia | Did not receive oxygen |
| Moderate | Clinical signs of non-severe pneumonia AND SpO<sub>2</sub>≥90% on room air | Did not receive oxygen |
| Severe | Clinical signs of severe pneumonia AND SpO<sub>2</sub>≤90% on room air; OR RR>30 breaths/min | Received oxygen (or told you they needed it, but it was not available) |
| Critical | ARDS; OR sepsis/septic shock; OR pulmonary embolism, acute coronary syndrome, acute stroke | Received invasive ventilation (or max available respiratory support) |

**Table S2.** 7T MRI sequence parameters. T1-MP2RAGE, T2-FLAIR, and T2-TSE images are analyzed for intracranial volume, white matter hyperintensity volume, and hippocampal subfield volume.

|  | T1-MP2RAGE | T2-FLAIR | T2-TSE |
| --- | --- | --- | --- |
| Resolution (mm) | 0.55 x 0.55 x 0.55 | 0.75 x 0.75 x 1.5 | 0.375 x 0.375 x 1.5 |
| Echo Time (ms) | 2.22 | 99 | 61 |
| Inversion Time (ms) | 800 & 2500 | 2900 | - |
| Repetition Time (ms) | 6000 | 14000 | 10060 |
| Acceleration Factor | 2 | 2 | 2 |
| Acquisition Time (min) | 12:44 | 11:28 | 3:53 |

